# Extraction of Human Phenotype Ontology (HPO) Concepts from Clinical Notes Utilizing Large Language Models (LLM) with Model Context Protocol (MCP)

**DOI:** 10.64898/2026.05.23.26353963

**Authors:** Michael Larsen, Ian M. Campbell, Lori A. Orlando, Peter Robinson, Nephi A. Walton

**Affiliations:** Division of Medical Genetics, University of Utah Health, Salt Lake City, UT, USA; Division of Human Genetics, Children’s Hospital of Philadelphia, Philadelphia, PA, USA; Department of Medicine, Wake Forest University School of Medicine, NC, USA; Rahel Hirsch Center for Translational Medicine, Berlin Institute of Health at Charité, Berlin, Germany; The Jackson Laboratory for Genomic Medicine, Farmington, CT, USA; Center for Artificial Intelligence Research, Wake Forest University School of Medicine, NC, USA; Department of Epidemiology and Prevention, Wake Forest University School of Medicine, NC, USA

**Keywords:** Clinical Genomics, Artificial Intelligence, Natural Language Processing, Large Language Models, Human Phenotype Ontology

## Abstract

**Background:** Accurate extraction of Human Phenotype Ontology (HPO) terms from clinical notes is essential for variant prioritization and genetic diagnosis. Large language models (LLMs) often struggle to balance precision, hallucination avoidance, and ontology mapping accuracy, and prior work has shown that retrieval-based grounding can improve performance for individual models. We hypothesized that real-time ontology grounding through external tools would improve these metrics across heterogeneous LLMs, and we evaluated the Model Context Protocol (MCP), a standardized open framework for integrating external tools, as a vendor-agnostic mechanism for delivering such grounding.

**Methods:** Five LLMs (Claude Sonnet 4.5, GPT-5.1, Gemini 2.5 Pro, Grok 4.1, and Qwen3 30B) extracted HPO terms from four synthetic clinical genetics notes under two conditions: baseline (“No Tools,” internal knowledge only) and tool-augmented (“With Tools”), with real-time HPO retrieval delivered through MCP for models with native support and through functionally equivalent native tool-calling interfaces otherwise. Each model performed ≥50 runs per note per condition (>2,000 total runs). Performance was evaluated using Precision, Recall, and F1-score. Outputs were manually adjudicated to classify mapping errors and hallucinations. Results were benchmarked against a commercial EHR-based HPO extraction tool.

**Results:** Tool augmentation significantly improved performance across all models. Mean aggregate F1-score increased from 0.46 (SD 0.22) in the baseline condition to 0.72 (SD 0.15) with tools (p < 0.001). Mapping Error Rate decreased from 40.9% to 7.8% (p < 0.001), and Precision increased from 56% to 90%. Performance gains were observed across all model families, including the open-weight Qwen3 model (F1 0.11→0.50). For inferred phenotypes, F1 improved from 0.20 to 0.34 (p < 0.001) without a significant increase in hallucination rate (p = 0.08). Compared with the commercial benchmark, tool-augmented LLMs achieved higher F1-scores and substantially greater recall for inferred phenotypes.

**Conclusions:** Real-time ontology grounding substantially improves HPO extraction across diverse LLMs by reducing mapping errors and enhancing phenotype inference. The Model Context Protocol provides a standardized, interoperable mechanism for delivering such grounding, supporting reproducible, vendor-agnostic deployment of clinical LLM pipelines in genomic medicine.

**40-Word Summary:** Real-time ontology grounding substantially improves Human Phenotype Ontology term extraction from clinical notes across diverse large language models. The Model Context Protocol provides a standardized, vendor-agnostic mechanism for delivering this grounding, supporting interoperable clinical AI deployments.

## INTRODUCTION

The Human Phenotype Ontology (HPO) is a standardized ontology that provides a structured and controlled set of terms for describing phenotypic abnormalities and clinical features encountered in human disease[1]. Terms from this ontology are essential to genetic laboratories for variant prioritization and for clinical research. Yet, they are currently manually extracted by clinical geneticists or genetic counselors and then provided to the laboratory for clinical diagnosis, a time consuming and inefficient process. To address these challenges, several studies have previously evaluated the efficiency of using traditional Natural Language Processing (NLP) methods to extract HPO terms, and more recently large language models (LLMs). LLMs are advanced artificial intelligence (AI) tools based on transformer architectures that are trained on large datasets to comprehend and generate text in a human-like manner[2]. Study outcomes show that LLMs tend to outperform traditional NLP methods but have problems with consistency[3,4]. Specifically, during HPO extraction, LLMs have been subject to hallucinations and inaccurate mapping to HPO terms[5,6].

However, LLM models are rapidly evolving and many commercially available LLMs incorporate different feature sets and reasoning capabilities. One recent advance is the Model Context Protocol (MCP)[7], an open-source standard introduced by Anthropic in November 2024 that standardizes secure two-way communication between LLMs and external data sources or tools, enabling AI applications to access real-time information without custom integration. By treating an ontology resource as a model-agnostic server, MCP enables LLMs across vendors to access and utilize information repositories, such as the HPO terms database, through a single shared interface.

Given the critical role of HPO terms in clinical genetic diagnosis and the expanding use cases for genomic medicine, there is an increasingly imperative need for rapid, scalable, and efficient HPO extraction processes. We, therefore, evaluated the performance of a broad variety of LLMs including, some of the most popular ones, such as those developed by Anthropic, OpenAI, Google, and xAI. In addition, because most healthcare environments restrict uploading clinical text to third-party APIs, we included Qwen3 as a locally deployable open-weight comparator, selected for its reasoning-style inference and access to MCP-enabled tools[8]. Lastly, given its clinical availability, we included the recently released LLM-based HPO extraction tool in Epic’s commercially available electronic health record (EHR) software[9]. We hypothesized that real-time HPO retrieval would improve extraction accuracy across heterogeneous LLMs, and we evaluated MCP as a vendor-agnostic mechanism for delivering such grounding.

## METHODS

### Overall study design

In this study we extracted HPO terms from four synthetic clinical notes using five different LLMs. Each synthetic clinical note was analyzed independently by each LLM across 50 repeated runs to account for variability. We employed a comparative study design to evaluate the impact of MCP tool augmentation on phenotype extraction: a baseline “No Tools” condition (internal knowledge only) versus a “Tool-Augmented” condition (real-time HPO database access).

### Large Language Model Selection

We selected five frontier LLMs that represent a diverse range of architectures, reasoning capabilities, and deployment modalities. To ensure a comparison of clinical reasoning, we exclusively selected models that support “Chain of Thought” (CoT) or equivalent reasoning modes. Commercial products included Claude Sonnet 4.5 (claude-sonnet-4-5-20250929; Anthropic) and Grok 4.1 Fast Reasoning (grok-4-1-fast-reasoning; xAI), both selected for their advanced reasoning performance and native support for MCP. We also included GPT-5.1 (gpt-5.1-2025-11-13; OpenAI) and Gemini 2.5 Pro (gemini-2.5-pro; Google) to evaluate performance using native function calling libraries. To assess feasibility of privacy-centric, local clinical pipelines, we also included Qwen3 30B (qwen3:30b-a3b), a 30-billion parameter open-weight model deployed locally via Ollama, to simulate performance on accessible, resource-constrained hardware.

### Clinical Note Creation

Four synthetic clinical genetics notes, modeled on real patient encounters, were independently authored by two clinical geneticists. Each note adhered to a standard clinical documentation format consistent with routine genetics practice and was not structured or altered to facilitate automated phenotype extraction. The notes included demographic data, laboratory values, anthropometric measurements, and quantitative clinical details to permit diagnostic inference in cases where key phenotypes were not explicitly stated in narrative form. Each note was designed to include specific “distractor” elements, including phenotypic information embedded within the family history and the review of systems using explicit negations. This design enabled the evaluation of model performance for distinguishing patient-specific phenotypes from those attributable to relatives and for correctly interpreting negated clinical findings.

### MCP Integration Architectures

To accommodate varying levels of support for MCP, we implemented three integration architectures designed to achieve functional similarity across platforms. Central to all architectures was the search_hpo_terms tool, which allowed models to query the HPO using natural language or fuzzy terms.

For models with native MCP support (Claude and Grok), we deployed a remote MCP server compliant with the JSON-RPC 2.0 standard over Server-Sent Events (SSE)[10]. The server configuration was passed directly in the API request headers, allowing these models to natively discover the search_hpo_terms tool. For the local Qwen3 model, we developed a custom Python MCP Client middleware that bridged the Ollama inference server with the same remote MCP server. In both setups, the remote server executed a rank-scored search against the HPO database, prioritizing exact ID and label matches over synonyms or partial substrings, returning the top 15 results to the model’s reasoning stream.

For models lacking native MCP support at the time of analysis (GPT-5.1 and Gemini 2.5 Pro), we simulated the protocol using native Function Calling APIs. We injected a JSON schema definition for search_hpo_terms into the system prompt, mirroring the schema used by the MCP server. When a model emitted a tool call, a client-side interceptor executed a local Python function that replicated the remote server’s logic against a local HPO JSON snapshot. This local function utilized an identical ranking algorithm, and returned the top 15 results. This approach ensured that the information retrieval capabilities and search granularity were consistent with the MCP-enabled conditions.

### Model Performance Measures

Measures included True Positives (TP), True negatives (TN), False positives (FP), and False Negatives (FN). TPs were extracted terms that matched the reference phenotype set (from the clinical note) via exact or synonymous mapping, hierarchical consistency (parent/child relationships), or clinical equivalence. False Positives were all other extracted phenotypes. FNs were reference phenotypes present in the clinical note but not identified during extraction; and TNs were the absence of both an extracted and referenced phenotype.

All extracted outputs were manually adjudicated by a clinical geneticist against the source note and labeled as correct or incorrect. Incorrect outputs were further classified as either mapping errors or hallucinations. Mapping errors occurred when a phenotype supported by the note was identified but mapped to an incorrect or non-existent HPO identifier; these outputs were counted as false positives for metric computation, and the corresponding reference phenotype was counted as a false negative if not otherwise captured correctly. Hallucinations were defined as any extracted phenotype not explicitly supported by the clinical note, laboratory data, or imaging; these were always counted as false positives. Hallucinations included fabricated findings, misattribution of family history to the patient, reversal of negated findings, and unsupported diagnostic inferences (e.g., mapping “systolic murmur” to a specific congenital heart defect without confirmatory imaging).

### Inferred vs Direct Classification

We stratified extraction performance based on whether the phenotype was explicitly stated in the text (“Direct”) or required clinical synthesis of data points (“Inferred”). Direct Phenotypes were defined as those explicitly stated in the clinical text, including standard medical terminology and common synonyms. Inferred Phenotypes were defined as those requiring the synthesis of multiple data points or the interpretation of descriptive findings to derive a feature not explicitly named. Each note and the target terms are available in the supplementary materials.

### Cost Assessment

To evaluate the operational feasibility of deploying these architectures in healthcare settings, we logged granular token usage (input, output, and reasoning tokens) for every iteration. We performed a comparative cost analysis for commercial models based on standard API pricing models (as of December 2025) to quantify the economic trade-off between the increased token overhead of tool use and the resulting improvements in data integrity. Notably, these figures reflect baseline performance; explicit optimization of the pipeline for cost-efficiency was considered outside the scope of this initial study. For the local open-weight model (Qwen), where direct API costs are not applicable, we measured token throughput and total generation count on a representative subset of runs.

### Outcomes

Model performance was evaluated using F1-score (primary outcome) and Precision, and Recall (secondary outcomes). Precision was defined as TP / (TP + FP), Recall as TP / (TP + FN), and F1 as 2 × (Precision × Recall) / (Precision + Recall).

### Statistical Analysis

Quantitative analysis was performed on an aggregated dataset comprising over 2,000 total extraction runs. This dataset was generated by performing at least 50 independent iterations for each of the four clinical notes across all five models and both experimental conditions. We calculated mean performance metrics for each model under both conditions. To evaluate the statistical significance of improvements conferred by tool integration, we compared the “No Tools” and “Tool-Augmented” conditions using Python (scipy.stats)[11]. Given that phenotype extraction performance metrics across stochastic iterations often violate normality assumptions, we employed a dual-testing framework to ensure robustness, utilizing Welch’s t-test[12] (two-sided, assuming unequal variances) to compare mean performance and the non-parametric Mann-Whitney U[13] test to validate results against non-normal distributions. Statistical significance was defined as p < 0.05.

### Comparison with EPIC

To benchmark performance against a commercially available clinical tool, we evaluated the new generative AI-driven phenotype extraction functionality native to the Epic EHR platform (Epic Systems Corporation, Verona, WI) [9]. The same four synthetic notes were instantiated within a secure, non-production EHR testing environment. To assess intra-system consistency, each of the four notes were entered into five distinct simulated patient charts, yielding a total of 20 simulated patient records. The clinical text was documented as standard ambulatory progress notes to simulate routine data entry. HPO terms were then generated using Epic’s integrated AI phenotyping tool. The resulting outputs were exported and subjected to the identical manual adjudication processed used for the LLM outputs. All software tools, synthetic clinical notes, and evaluation scripts used in this study are available via GitHub at: [https://github.com/clinical-mcp/pheno-extract-ai]. The released materials correspond to a versioned snapshot of the repository (v1.0.0) and are provided under an academic-use–only license. Use of these resources is restricted to non-commercial research and educational purposes, and appropriate citation of this work is required. Commercial use, redistribution, or derivative commercial applications are not permitted without prior written authorization from the authors. Interested commercial users should contact the corresponding authors for licensing inquiries.

The Human Phenotype Ontology (HPO) Model Context Protocol (MCP) server used in this study is available at: [https://github.com/clinical-mcp/hpo_mcp]. The HPO MCP repository includes instructions for reproducible local deployment using Docker. A publicly accessible hosted instance is also described in the repository README for convenience; however, reproduction of the study methods does not depend on continued availability of that endpoint, as the full containerized implementation is provided in the repository.

## RESULTS

### Overall Efficacy of Tool Augmentation

Real-time HPO retrieval, delivered through MCP where natively supported and through equivalent tool-calling interfaces otherwise, yielded statistically significant improvements in the primary outcome across all evaluated models. In the baseline “No Tools” condition, models achieved a mean aggregate F1-score of 0.46 (SD 0.22). Enabling real-time HPO access raised the mean F1-score to 0.72 (SD 0.15), a statistically significant improvement in extraction performance (p < 0.001) with a large effect size (Cohen’s d > 0.8). This improvement was driven by dual gains in sensitivity and mapping reliability. The Mapping Error Rate decreased from 40.9% in the baseline condition to 7.8% with tools (p < 0.001). Aggregate Precision increased from 56% to 90%, indicating that knowledge grounding reduced incorrect ontology mappings and improved semantic accuracy. Model comparisons for F1, recall, and precision are visualized in Figure 2. Mapping error rates and hallucination rates for each of the models are summarized in Figure 3.

**Figure 1.**
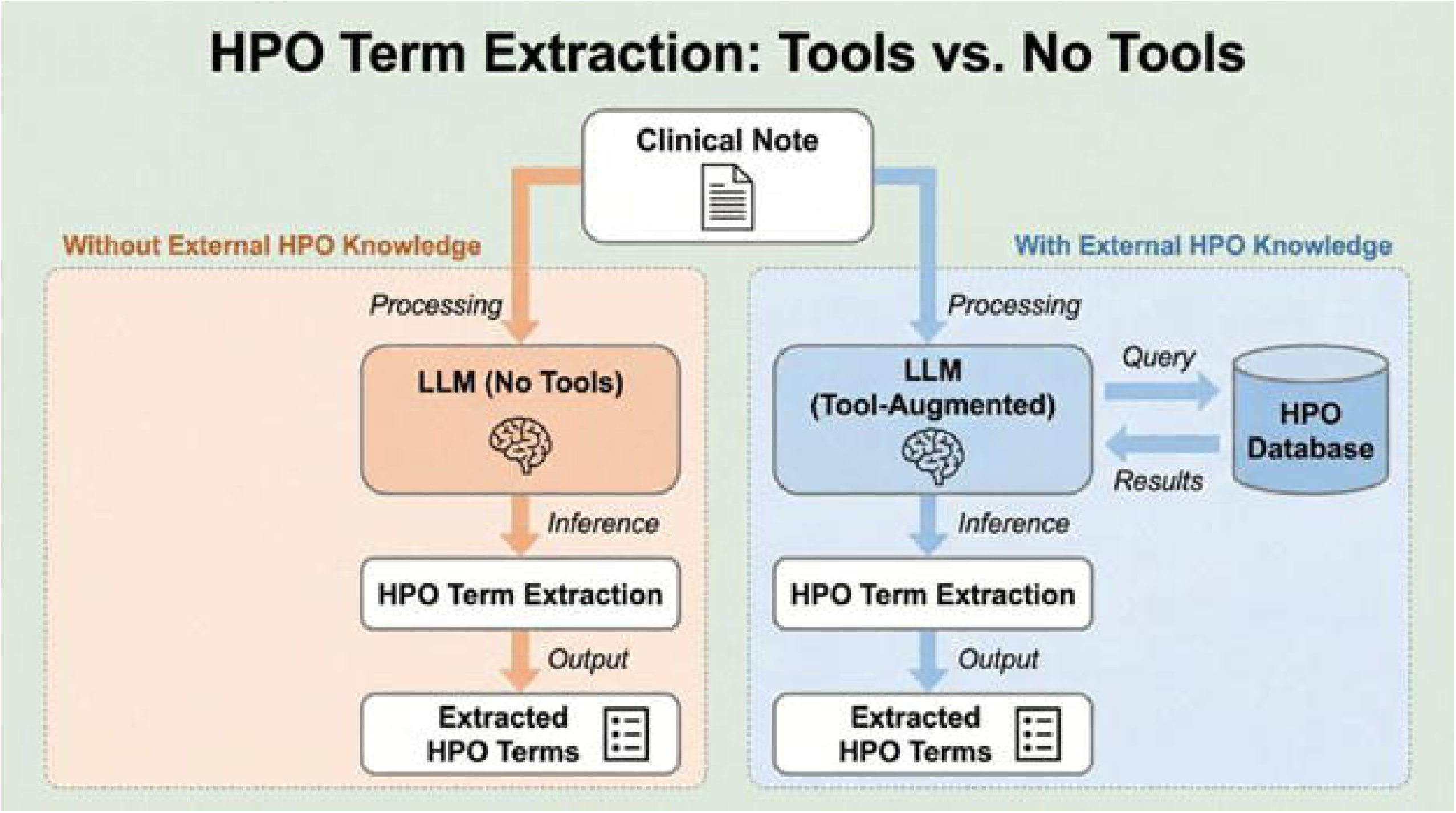
Study workflow for HPO term extraction with and without external ontology access. Legend: Schematic of the two experimental conditions used for HPO term extraction from clinical notes. In the baseline “No Tools” condition, the LLM processes the clinical note using internal model knowledge alone and directly outputs extracted HPO terms. In the tool-augmented condition, the LLM queries an external HPO database during inference and uses returned ontology results to support phenotype identification and term mapping before producing extracted HPO terms. This figure illustrates the architectural distinction underlying the comparative evaluation. **Alt text:** Diagram comparing 2 workflows for HPO extraction from a clinical note. On the left, an LLM without tools processes the note and outputs HPO terms directly. On the right, a tool-augmented LLM queries an external HPO database during processing and then outputs HPO terms.

**Figure 2.**
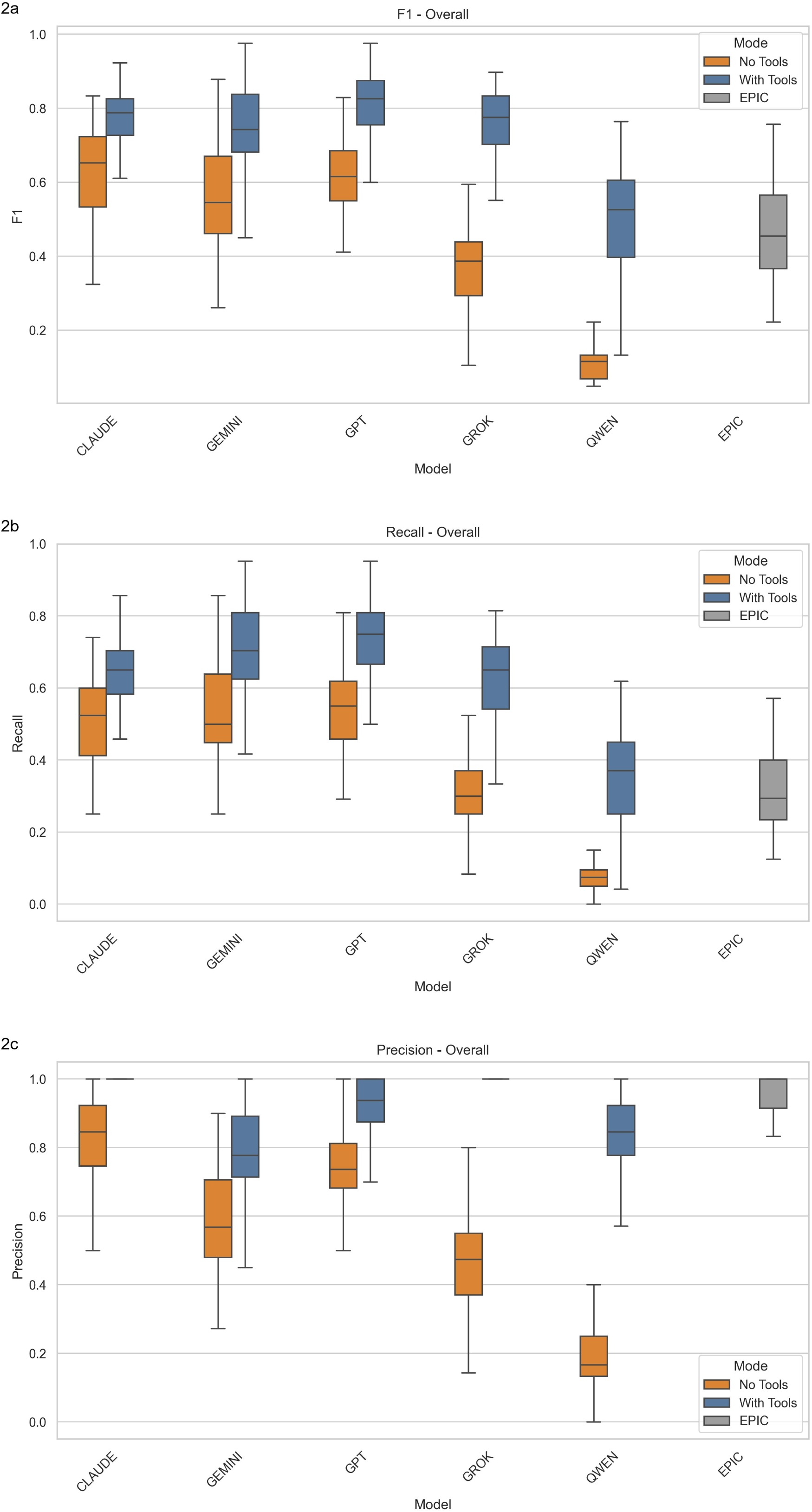
Overall extraction performance across models and tool-use conditions. **Legend:** Comparison of overall extraction performance across evaluated models under baseline (“No Tools”) and tool-augmented (“With Tools”) conditions, with the Epic EHR phenotype tool shown as a comparator. (A) Overall F1-score. (B) Overall recall. (C) Overall precision. Box plots summarize distributions across repeated extraction runs for each model and condition. Across all models, ontology-grounded tool use improved overall extraction performance, primarily through gains in recall and F1-score while maintaining or improving precision. Alt text: Three box-plot panels comparing overall F1, recall, and precision across models with and without tools, plus an Epic comparator. Tool-augmented conditions generally show higher performance than no-tools conditions..

**Figure 3.**
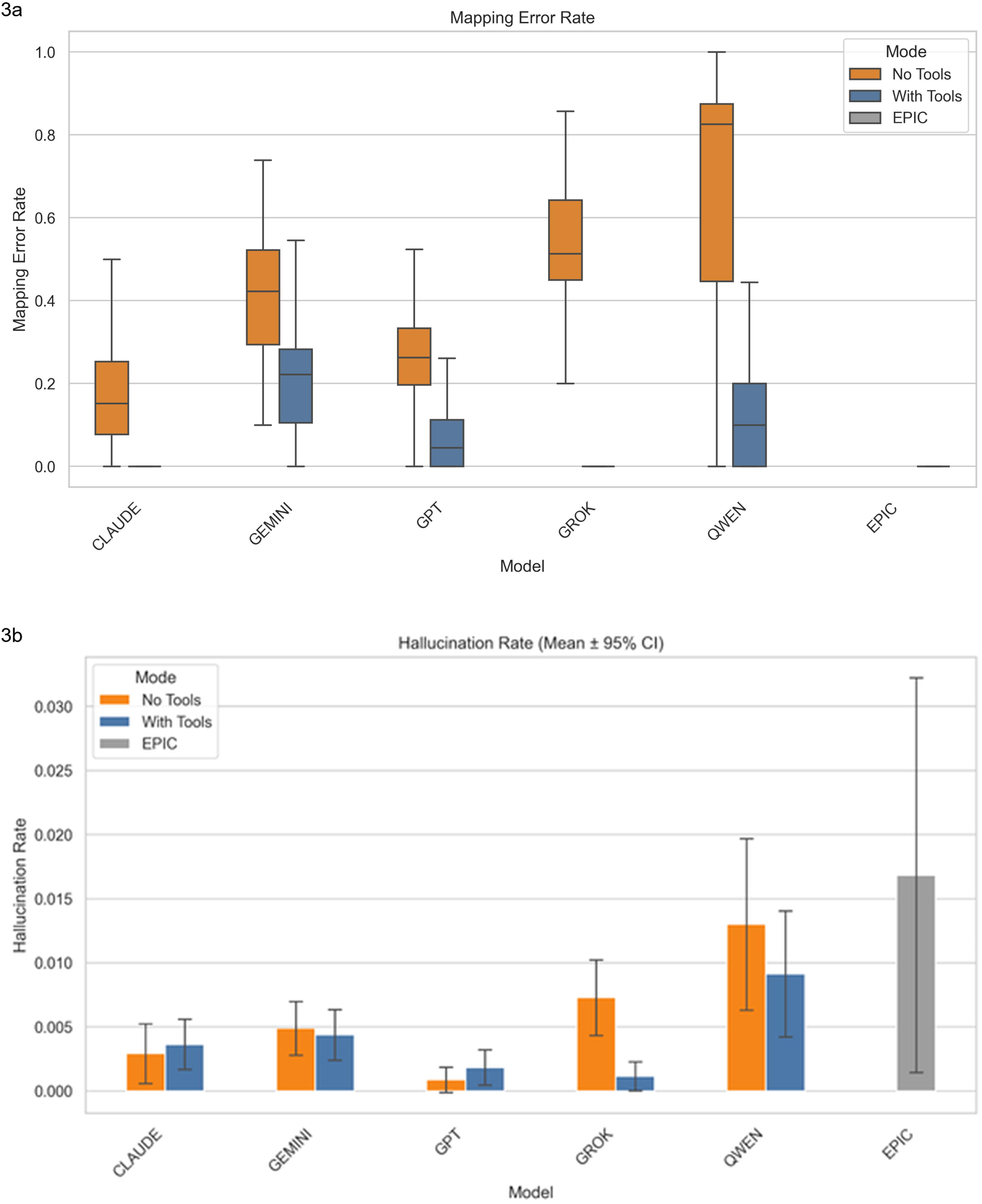
Error profiles across models and tool-use conditions. **Legend**: Comparison of extraction error profiles across evaluated models under baseline (“No Tools”) and tool-augmented (“With Tools”) conditions, with the Epic EHR phenotype tool shown as a comparator. (A) Mapping error rate. (B) Hallucination rate shown as mean ± 95% confidence interval. Tool augmentation substantially reduced mapping errors across models, while hallucination rates remained low overall and did not show a marked increase with tool use. Alt text: Two panels comparing mapping error rates and hallucination rates across models with and without tools, plus Epic as comparator. Mapping errors are much lower with tools, while hallucination rates remain low overall.

### Model-Specific Performance

Performance gains were consistent across all five evaluated models (Table 1). GPT-5.1 achieved the highest absolute performance in the tool-enabled condition, with its F1-score increasing from 0.62 to 0.81 (p < 0.001). Claude Sonnet 4.5 was the most reliable ontology mapper; while its F1-score improved from 0.62 to 0.78, its notable achievement was a near-total elimination of mapping errors (19.2% to 0.04%), effectively achieving perfect adherence to HPO. Grok 4.1 exhibited the largest relative gain; in the baseline condition, it struggled (F1 0.36), but tool access more than doubled its F1-score to 0.76 (p < 0.001). Finally, the local open-weight model, Qwen 30B, demonstrated that MCP use can redeem smaller architectures. While its baseline performance was non-viable (F1 0.11), the addition of the MCP client raised its F1-score to 0.50 (p < 0.001), rendering it functionally useful for basic extraction tasks.

**Table 1.**
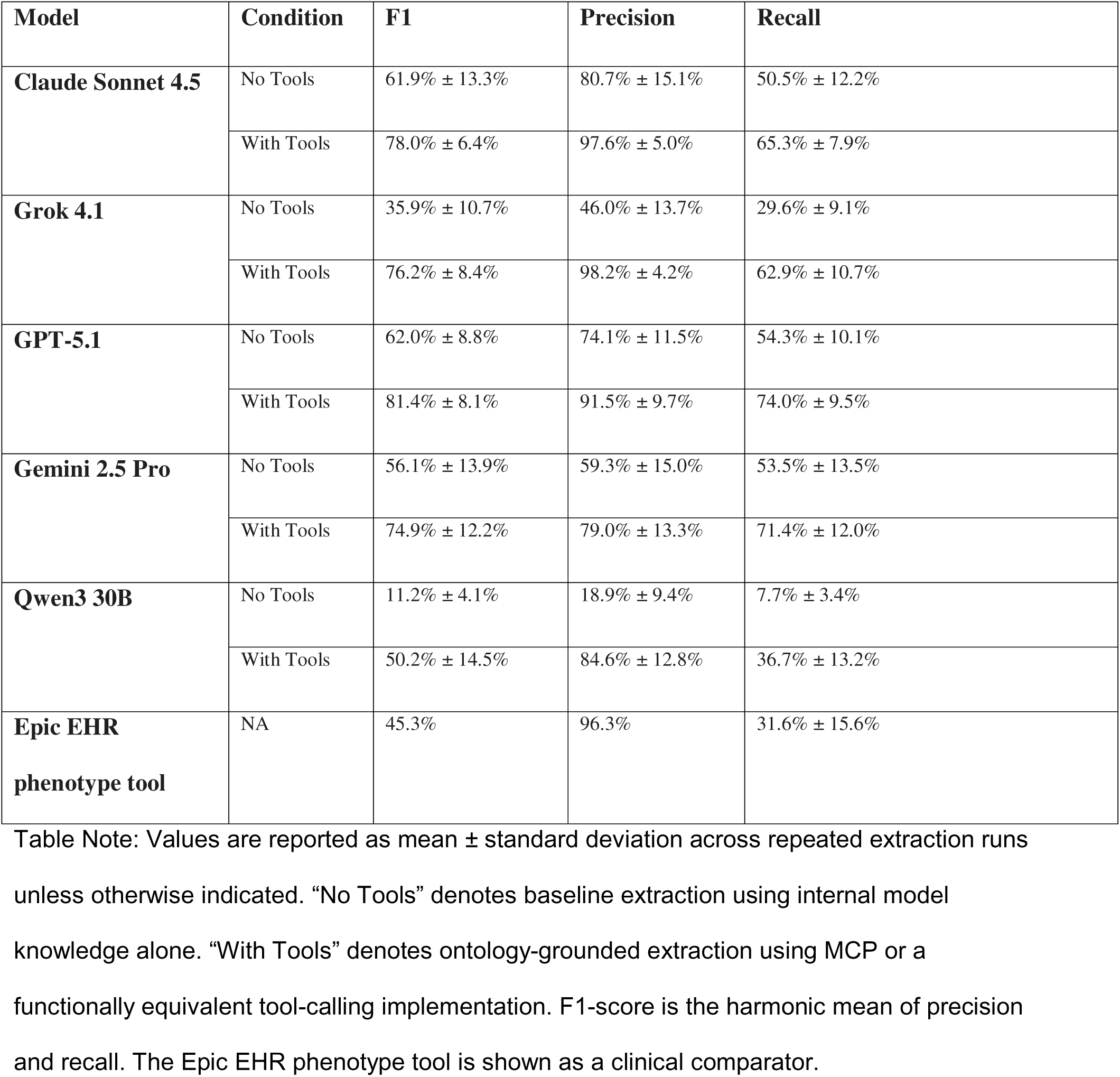
Overall HPO extraction performance by model and tool-use condition.

| Model | Condition | F1 | Precision | Recall |
| --- | --- | --- | --- | --- |
| **Claude Sonnet 4.5** | No Tools | 61.9% ± 13.3% | 80.7% ± 15.1% | 50.5% ± 12.2% |
| | With Tools | 78.0% ± 6.4% | 97.6% ± 5.0% | 65.3% ± 7.9% |
| **Grok 4.1** | No Tools | 35.9% ± 10.7% | 46.0% ± 13.7% | 29.6% ± 9.1% |
| | With Tools | 76.2% ± 8.4% | 98.2% ± 4.2% | 62.9% ± 10.7% |
| **GPT-5.1** | No Tools | 62.0% ± 8.8% | 74.1% ± 11.5% | 54.3% ± 10.1% |
| | With Tools | 81.4% ± 8.1% | 91.5% ± 9.7% | 74.0% ± 9.5% |
| **Gemini 2.5 Pro** | No Tools | 56.1% ± 13.9% | 59.3% ± 15.0% | 53.5% ± 13.5% |
| | With Tools | 74.9% ± 12.2% | 79.0% ± 13.3% | 71.4% ± 12.0% |
| **Qwen3 30B** | No Tools | 11.2% ± 4.1% | 18.9% ± 9.4% | 7.7% ± 3.4% |
| | With Tools | 50.2% ± 14.5% | 84.6% ± 12.8% | 36.7% ± 13.2% |
| **Epic EHR**<br>**phenotype tool** | NA | 45.3% | 96.3% | 31.6% ± 15.6% |

Table Note: Values are reported as mean ± standard deviation across repeated extraction runs unless otherwise indicated. “No Tools” denotes baseline extraction using internal model knowledge alone. “With Tools” denotes ontology-grounded extraction using MCP or a functionally equivalent tool-calling implementation. F1-score is the harmonic mean of precision and recall. The Epic EHR phenotype tool is shown as a clinical comparator.

### Direct vs. Inferred Phenotypes

We stratified extraction performance based on whether the phenotype was explicitly stated in the text (“Direct”) or required clinical synthesis (“Inferred”) (Table 2). Across all evaluated LLMs, direct phenotype extraction consistently outperformed inferred phenotype extraction in both experimental conditions. Mean direct F1 increased from 0.50 in the baseline condition to 0.76 with tool augmentation, while mean inferred F1 improved from 0.20 to 0.34 (p < 0.001). Tool augmentation improved inferred phenotype extraction for every model tested. Gemini 2.5 Pro demonstrated the strongest performance on inferred phenotypes in the tool-enabled condition (F1 0.53), while GPT-5.1 achieved the highest direct phenotype performance (F1 0.85). The largest gains in inferred phenotype extraction were observed for Grok 4.1 (0.13 to 0.38) and Gemini 2.5 Pro (0.31 to 0.53). Importantly, the improved recovery of inferred phenotypes did not coincide with a significant increase in hallucination rate (p = 0.08), suggesting that ontology-grounded tool use improves clinical synthesis without substantially increasing fabricated findings.

**Table 2:**
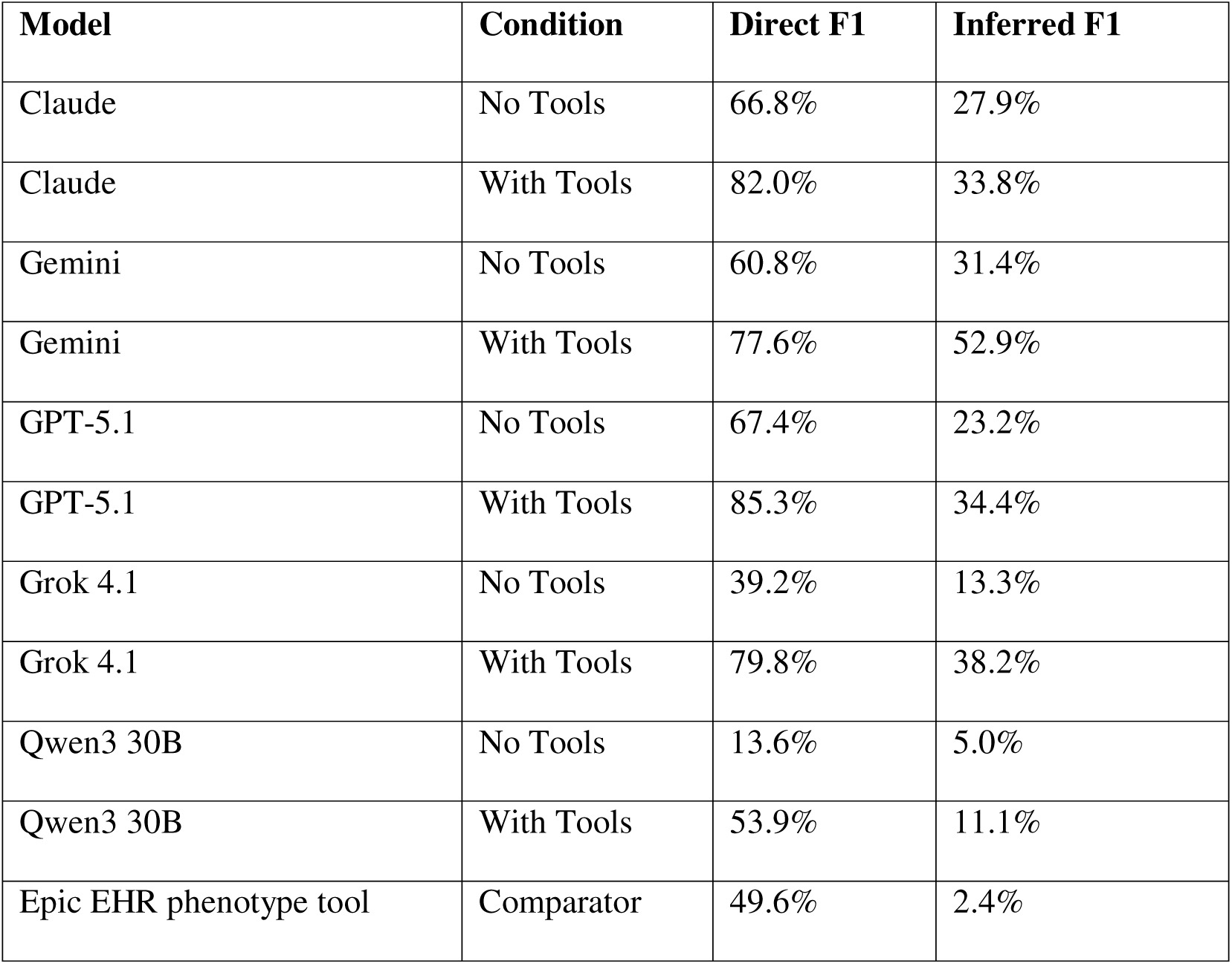
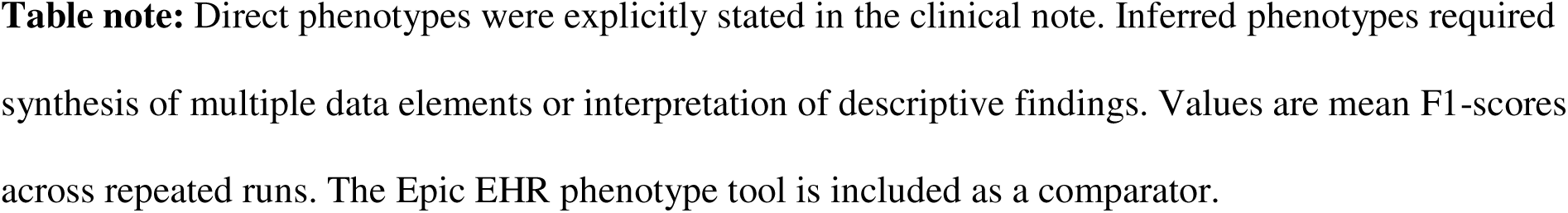
Extraction performance for direct versus inferred phenotypes by model and tool-use condition.

| Model | Condition | Direct F1 | Inferred F1 |
| --- | --- | --- | --- |
| Claude | No Tools | 66.8% | 27.9% |
| Claude | With Tools | 82.0% | 33.8% |
| Gemini | No Tools | 60.8% | 31.4% |
| Gemini | With Tools | 77.6% | 52.9% |
| GPT-5.1 | No Tools | 67.4% | 23.2% |
| GPT-5.1 | With Tools | 85.3% | 34.4% |
| Grok 4.1 | No Tools | 39.2% | 13.3% |
| Grok 4.1 | With Tools | 79.8% | 38.2% |
| Qwen3 30B | No Tools | 13.6% | 5.0% |
| Qwen3 30B | With Tools | 53.9% | 11.1% |
| Epic EHR phenotype tool | Comparator | 49.6% | 2.4% |

**Table note:** Direct phenotypes were explicitly stated in the clinical note. Inferred phenotypes required synthesis of multiple data elements or interpretation of descriptive findings. Values are mean F1-scores across repeated runs. The Epic EHR phenotype tool is included as a comparator.

To illustrate this distinction at the note level, Figure 4 shows term detection rates for one representative simulated clinical genetics note, with each phenotype annotated as direct or inferred. Consistent with the aggregate findings in Table 2, directly stated phenotypes were generally detected more reliably across models and conditions, whereas inferred phenotypes showed greater variability and remained more dependent on model choice and tool augmentation. This note-level example provides a concrete demonstration of the performance gap summarized in Table 2 and highlights the persistent challenge of extracting phenotypes that require synthesis rather than explicit text matching.

**Figure 4.** Representative note-level term detection rates for direct and inferred phenotypes. Term detection rates are shown for phenotypes extracted from one representative simulated clinical genetics note across models and tool-use conditions. The left annotation bar indicates whether each phenotype was classified as direct or inferred. Darker shading indicates higher detection frequency across repeated runs. This example illustrates the greater consistency of extraction for direct phenotypes and the higher variability observed for inferred phenotypes. **Alt Text:** Heatmap-style bar chart showing term detection rates for various phenotypes extracted from a single simulated clinical genetics note. Phenotypes are listed vertically, with detection rates across different models and tool-use conditions displayed horizontally via bars or cells. A left-side annotation bar labels each phenotype as either “direct” or “inferred.” Darker shading represents higher detection frequency across multiple repeated runs. Direct phenotypes generally show more consistent (less variable) detection rates, while inferred phenotypes exhibit greater variability and inconsistency.

#### Cost Assessment

As detailed in Table 3, the integration of MCP tools resulted in an expected increase in both mean token utilization and associated computational costs across all evaluated commercial models. This increase reflects the inherent overhead required for tool-calling sequences and expanded context processing. These figures represent the unoptimized, baseline execution of the extraction pipeline to establish a foundational performance floor.

**Table 3.** Mean token utilization and estimated per-run cost by model and tool-use condition.

| Model | Without Tools |  | With Tools |  |
| --- | --- | --- | --- | --- |
|  | Mean Cost (USD) | Mean Tokens | Mean Cost (USD) | Mean Tokens |
| Claude Sonnet 4.5 | 0.081 | 7508 | 0.178 | 44895 |
| Grok 4.1 | 0.002 | 5877 | 0.006 | 21163 |
| GPT-5.1 | 0.038 | 5655 | 0.083 | 14213 |
| Gemini 2.5 Pro | 0.006 | 5463 | 0.0345 | 20542 |
| Qwen3 30B | N/A | 30556 | N/A | 59696 |

**Table note:** Costs reflect unoptimized baseline execution using standard commercial API pricing as of December 2025. Token counts represent mean total tokens consumed per run. Qwen3 30B was deployed locally and therefore has no direct API cost.

### Comparison with Commercial Benchmarks

To contextualize these findings, we compared the LLMs against the phenotype extraction utility currently available in the Epic electronic medical record. The Epic tool demonstrated high precision but notably low recall. It successfully captured “Direct” phenotypes but failed to capture “Inferred” phenotypes, achieving an accuracy of only 2.4% on terms requiring synthesis of laboratory or imaging data. In contrast, the tool-augmented LLMs achieved up to 45% accuracy on these same inferred terms. This resulted in the Epic tool having a significantly lower overall F1-score compared to the tool-augmented LLMs, highlighting a clear divergence in utility between strict, rule-based extraction and generative reasoning.

## DISCUSSION

HPO terms are a critical component of implementing genomic medicine at scale, yet are rarely recorded in the medical record. In this study we report two related but distinct findings. Empirically, real-time HPO retrieval through external tools substantially improved phenotype extraction across all five evaluated LLMs. Architecturally, the same HPO retrieval logic delivered these gains across heterogeneous model families—including a locally hosted open-weight model—using a single, standardized interface, suggesting that MCP can serve as a vendor-agnostic foundation for clinical AI tooling.

In the baseline “No Tools” condition, even frontier reasoning models frequently output non-existent ontology terms or fail to map recognized concepts to their HPO identifiers. By enabling tools via an MCP server, we effectively decoupled the reasoning process (identification of a symptom in a text) from the retrieval process (producing the appropriate HPO term). This led to an improvement in both recall and precision across all tested models for all notes.

The comparison with the commercial benchmark (Epic EHR) highlights differences in term extraction. The Epic utility demonstrated high precision for explicitly documented phenotypes but lower recall relative to tool-augmented LLMs, particularly for inferred phenotypes requiring clinical synthesis. In contrast, MCP-augmented LLMs achieved higher overall recall and F1-scores, reflecting improved capture of both directly stated and inferred phenotypes. This distinction likely reflects differences in calibration strategy. A conservative extraction approach may be advantageous when processing large volumes of longitudinal clinical data across the medical record. In contrast, consult-level rare disease evaluation may benefit from a more recall-sensitive approach capable of deeper phenotype synthesis. Ontology-grounded LLM pipelines may therefore provide complementary value depending on clinical context. Because HPO terms directly inform variant prioritization and cohort matching workflows, improvements in extraction fidelity may have downstream implications for genomic analysis pipelines. As illustrated in Figure 4, a residual challenge was consistent synthesis of clinical information into inferred phenotype labels.

While previous studies have utilized custom Python functions or retrieval-augmented generation (RAG) to improve extraction [14–17], establishing the general benefit of ontology grounding, this study extends those findings by evaluating the Model Context Protocol as a standardized architectural interface for delivering that grounding. The principal advantage of MCP lies in its interoperability. By treating the HPO database as a modular, version-controlled server, the same ontology tool can be deployed across heterogeneous model families, ranging from cloud-based frontier systems to locally hosted open-weight models, without model-specific refactoring.

This architectural separation of reasoning (the model) from ontology validation (the server) offers practical infrastructure advantages. Healthcare systems can update or version the ontology server independently of model selection, and institutions may deploy local MCP instances alongside locally hosted models when data residency or privacy constraints require it. Such decoupling enables clinical AI pipelines to remain stable and vendor-agnostic despite rapid turnover in the LLM ecosystem. To facilitate reproducibility and independent validation, the HPO MCP server used in this study has been released as an open-source repository with containerized deployment instructions. The repository documents access to a public hosted instance for convenience, but reproducibility does not depend on that hosted endpoint. Repository preparation and deployment documentation were completed in coordination with collaborators at The Jackson Laboratory.

We therefore suggest that MCP, or functionally equivalent open standards, represents a viable specification for clinical AI tooling where reproducibility, consistency, and long-term maintainability are required.

While our cost assessment (Table 3) demonstrates an increased computational expense associated with tool augmentation, it is important to emphasize that these runs were not optimized for token efficiency. The primary objective of this study was to evaluate baseline extraction fidelity and ontology adherence; therefore, cost optimization was considered outside of our current scope and we are strictly reporting unoptimized utilization. It is highly likely that routine optimization strategies would yield improved operational costs. Future studies specifically designed to directly compare the economic viability and cost-to-benefit ratios of these models will be necessary to guide high-throughput clinical deployment. This study has limitations. First, the synthetic clinical notes were modeled on high-quality documentation common in academic clinical genetics, which tends to be more detailed and phenotypically rich than notes from primary care or other specialties. Therefore, the generalizability of our findings to sparse or highly unstructured community health records remains to be tested. Second, our evaluation was restricted to five generative AI models. While these represent the current state-of-the-art, the rapid pace of AI development means newer models may exhibit different behaviors.

Third, while we harmonized system prompts, intrinsic differences in model training data and safety alignment (e.g., refusal triggers) may have influenced comparative performance. Finally, our “Gold Standard” for recall was based on expert human annotation; however, in clinical practice, inter-provider agreement on HPO term selection is known to be imperfect, introducing an element of subjectivity to the “ground truth.”

## CONCLUSION

Grounding generative AI models with real-time ontology retrieval substantially improves HPO term extraction by reducing mapping errors and enabling reliable phenotype inference. These gains were observed consistently across multiple frontier and open-weight models. The Model Context Protocol provides a standardized, model-native mechanism for delivering such grounding; by separating reasoning from ontology validation, MCP offers a vendor-agnostic foundation for structured knowledge access that supports reproducibility and cross-model interoperability.

These findings support the integration of ontology-grounded retrieval mechanisms in clinical LLM pipelines, particularly in genomic medicine workflows where phenotype completeness and accuracy directly influence downstream analyses. Adoption of MCP, or functionally equivalent open standards, may facilitate scalable, vendor-agnostic deployment of phenotype extraction systems within clinical infrastructure.

## DATA AVAILABILITY

All software tools, synthetic clinical notes, gold-standard annotations, and evaluation scripts used in this study are publicly available via GitHub at https://github.com/clinical-mcp/pheno-extract-ai and https://github.com/clinical-mcp/hpo_mcp. The complete run-level result dataset and final curated annotations will be deposited in the Dryad Digital Repository upon publication, and a draft DOI will be reserved at that time.

